# Characteristics of HIV cure-related clinical trials affecting study participation: A choice-based conjoint analysis experiment

**DOI:** 10.64898/2026.10.03.26364635

**Authors:** Jerome T. Galea, Alicia Junker, Karah Y. Greene, Andrea N. Polonijo, Karine Dubé, Jeff Taylor, Christopher Christenson, Brandon Brown

## Abstract

Cash incentives are widely used in clinical research to support recruitment and retention, yet their ethical implications, particularly in high-risk settings such as HIV cure-related studies, remain inadequately understood. This study examined how key stakeholders perceive participant payment relative to other study characteristics when deciding whether to participate in HIV cure research. Using a choice-based conjoint (CBC) experiment, we assessed preferences among people living with HIV (PLWH), HIV researchers, and Institutional Review Board (IRB) members/bioethicists. Participants (N=88) completed eight choice tasks involving hypothetical HIV cure study scenarios varying across six attributes: incentive amount, invasiveness, requirement to pause HIV medications, time commitment, first-in-human status, and cost reimbursement.

Across all participants, payment amount emerged as the most influential attribute (29.3%), followed by procedure invasiveness (22.3%) and the requirement to pause HIV medication (17.6%). The highest hypothetical payment level ($200 per visit) was significantly preferred. However, subgroup analyses revealed important differences: PLWH prioritized payment most strongly, whereas researchers and IRB members/bioethicists placed greater emphasis on minimizing invasiveness. The requirement to pause HIV medication was a salient concern across groups, though relatively more influential for IRB members/bioethicists than for PLWH. Time commitment and ancillary cost reimbursement had comparatively lower impact on decision-making.

These findings highlight the central role of financial incentives in shaping willingness to participate in HIV cure research, raising ethical considerations regarding potential undue influence, especially in higher-risk studies. At the same time, stakeholders value minimizing procedural risks and disruptions to treatment. This study underscores the need for clearer guidance on appropriate payment practices and for balancing incentives with ethical safeguards, particularly in trials involving antiretroviral therapy interruption.

## INTRODUCTION

Incentives, defined as a type of “payment(s) (including money, gifts, and services) to research volunteers for participation in studies,” are a widespread practice in clinical research [1, 2, 3, 4, 5]). Among types of payment for research, incentive payments may be the most ethically problematic [6]. Despite differing views, cash payments are often considered necessary to ensure participation in research [7, 8, 9]), especially in high-risk trials [10, 11]). Research shows that incentives may encourage participation and aid in participant retention in both clinical and behavioral research [8, 12, 13, 14]. Still, to avoid undue inducement, incentives should be appropriate for participants. Federal law prohibits using payments to research participants to offset risks posed by the study procedures [15, 16]. However, participants may believe that incentives are a benefit of participating in research despite the risks and assume that the payment amount compensates for those risks [17, 18]. In doing so, ethical questions may arise regarding payment (i.e., whether participants are assuming risks solely for money), and, for this reason, the balance of risks and potential benefits of study participation must be carefully considered [19, 14, 20, 21].

Many study characteristics influence the amount of cash payment provided to study participants, including the study’s benefits, risks, burden, time commitments, budget, recommendations from the Institutional Review Board (IRB), and local regulations [8, 22, 23, 24]. In HIV-related research, additional factors may also affect incentive amounts due to inherent levels of risks related to certain procedures (e.g., lumbar punctures, pausing antiretroviral therapy [ART]). Additionally, some people experiencing disparities, such as low socioeconomic status, may be differentially motivated to participate [5, 23, 25, 19].

Although the use of cash incentives is widespread in clinical research, little is known about their effects on participation in HIV cure-related research [3, 26, 27, 28]. Cash payments (and other study-related incentives) are not universally tracked; investigators may not record participant payment amounts in research protocols, and payment amounts are rarely reported in published manuscripts [29]. To address this knowledge gap, we conducted a choice-based conjoint analysis (CBC) experiment to examine how key HIV cure research stakeholders perceive the amount of cash payments received by study participants relative to other study features when considering participation.

## METHODS

### Overview

This study was a collaboration among academic partners, the non-profit organization The HIV + Aging Research Project | PS (H+ARP-PS), and a study-specific Community Advisory Board (CAB) comprising 12 men and women in the United States, including people living with HIV (PLWH), HIV researchers, IRB members, and bioethicists. The CAB, H+ARP-PS, and academic partners met monthly during the two-year study to jointly inform the contents of the study materials (documents, instruments, recruitment materials) and oversee study implementation. The study was approved by the Institutional Review Board at the University of California, Riverside (IRB # HS-20-248), with other participating universities deferring to that approval. All research participants provided informed consent prior to taking part in the study procedures. We first conducted a nationwide survey of PLWH on their perceptions of payment, then we conducted qualitative interviews and focus groups with stakeholder groups [17, 18, 30]. Here, we report results from the CBC experiment examining how payment amounts and other study attributes affect the decision to participate in a hypothetical HIV cure-related study.

### Participants and Setting

Participants comprised three groups: PLWH, HIV researchers, and IRB members/bioethicists, who, given their backgrounds and experience, were most likely to have opinions about the characteristics of HIV cure research studies. PLWH were recruited purposively from existing personal and professional networks from the research team and CAB members, as well as via circulation of an IRB-approved study recruitment flyer circulated to community-based HIV care organizations and professional email listservs known to the research team members. HIV researchers were purposively identified by the research team for their known experience in HIV research (clinical, behavioral, or both), and IRB members/bioethicists were likewise recruited from existing professional networks.

### Development of the Choice-Based Conjoint Analysis (CBC) Experiment

CBC is a market-based, quantitative research method that replicates how consumers make choices among competing products by requiring them to make trade-offs between the products [31]. Consumers are first shown a selection of nearly identical, and often hypothetical but plausible, products with varying attributes (characteristics). Then they choose their preferred product from the options. In doing so, CBC measures the importance of individual product attributes on the consumer’s product choice. CBC has been used in numerous health-related studies and has successfully predicted real-world health choices, including influenza vaccination and colorectal cancer screening, when scale and preference heterogeneity are accounted for [32, 33]). A systematic review also found that choice-based experiments can produce reasonable predictions of health-related behaviors [34]). Specific to HIV, conjoint analysis has been used to successfully predict HIV treatment preferences under different medication scenarios [35], as well as to elucidate product attribute preferences for hypothetical HIV prevention interventions [36, 37]).

The accuracy of CBC in predicting consumers’ preferences for hypothetical products depends greatly on the plausibility of the product options. Therefore, when designing the hypothetical HIV cure studies for the present CBC experiment, it was essential to select attributes that are both real and common in HIV cure research. At the same time, the total number of hypothetical study attributes used in CBC should be limited, typically to six or fewer, to prevent cognitive overload, which can weaken data validity [38]). Since HIV cure research studies have many more than six attributes, we conducted a three-step process to arrive at the six most important attributes to include in our CBC experiments: 1. Attribute Generation; 2. Attribute Ranking; 3. Attribute Selection.

#### Attribute generation

Research staff (academic and H+ARP-PS staff) brainstormed potential study attributes, yielding a preliminary list of attributes commonly encountered in HIV cure research studies. Overlapping attributes (i.e., attributes that were similar or that measured the same concept) were combined to ensure mutual exclusivity.

#### Attribute ranking

Next, the CAB reviewed the preliminary list of attributes and was invited to add any new attributes not on the original list, arriving at a final list of 17 attributes, which they individually rank-ordered according to their impact on deciding whether to participate in a HIV clinical research study (**Table 1**). CAB members were provided the following instructions prior to rank-ordering the attributes:

> Below is a list of possible characteristics of high-risk HIV clinical research studies that may affect your willingness to participate in the study. Please tell us how important these characteristics would be to your willingness to participate in high-risk HIV clinical research by sorting this list from most important to least important. “Drag and drop” this list so that the characteristics most important to you are at the top of the list and the characteristics least important to you are at the bottom of the list.

**Table 1:** CAB’s rank order of HIV potential cure-related study attributes (1=most important)

| Rank order | Potential HIV cure-related study attributes |
| --- | --- |
| 1 | The invasiveness of the study procedures (e.g., simple blood draws vs. spinal taps) |
| 2 | The overall duration of the study (e.g., days/weeks/months/years) |
| 3 | Whether antiretroviral (HIV medication) interruption is required |
| 4 | Whether study-related, out-of-pocket costs (e.g., parking, bus fare) are reimbursed |
| 5 | Whether a “washout” period (discontinuing current HIV treatment) is required before being able to participate |
| 6 | The number of clinic visits |
| 7 | The phase of the clinical trial (e.g., “first in humans”, Phase 1) |
| 8 | How the cash incentive is paid (e.g., gift card, direct cash payment) |
| 9 | The amount of cash incentive paid |
| 10 | Whether there are additional health care services provided (unrelated to the study itself) |
| 11 | The length of each study visit |
| 12 | Whether the study becomes more or less risky over time (e.g., due to stopping HIV meds) |
| 13 | Whether an overnight hospital stay is required |
| 14 | Whether there are possible risks to your partner as a result of study participation |
| 15 | Whether you'd have access to the intervention (if effective) after the study |
| 16 | Whether abstinence from sex is required (e.g., during a time when HIV meds are stopped) |
| 17 | Whether there is a restriction on simultaneous participation in a different study |

#### Attribute selection

To arrive at the final list of the six most important study attributes for the CBC experiment, we reviewed the attributes in **Figure 1** to ensure each was unique. For example, two preliminary attributes, “overall duration of the study” and “the number of study visits,” were combined into the new attribute, “time commitment.” Likewise, “whether antiretroviral (HIV medication) interruption is required” and “whether a washout period (discontinuing current HIV treatment) is required” were combined into the new attribute “Must stop HIV medications to participate.” Consistent with IRBs’ mandate to review participant payment amounts and our interest to specifically understand how research payments affect decisions to participate in HIV cure research, “amount of incentive” was included as one of the final six attributes despite its lower ranking (9^th^ place in Table 1) by the CAB.

**Figure 1:**
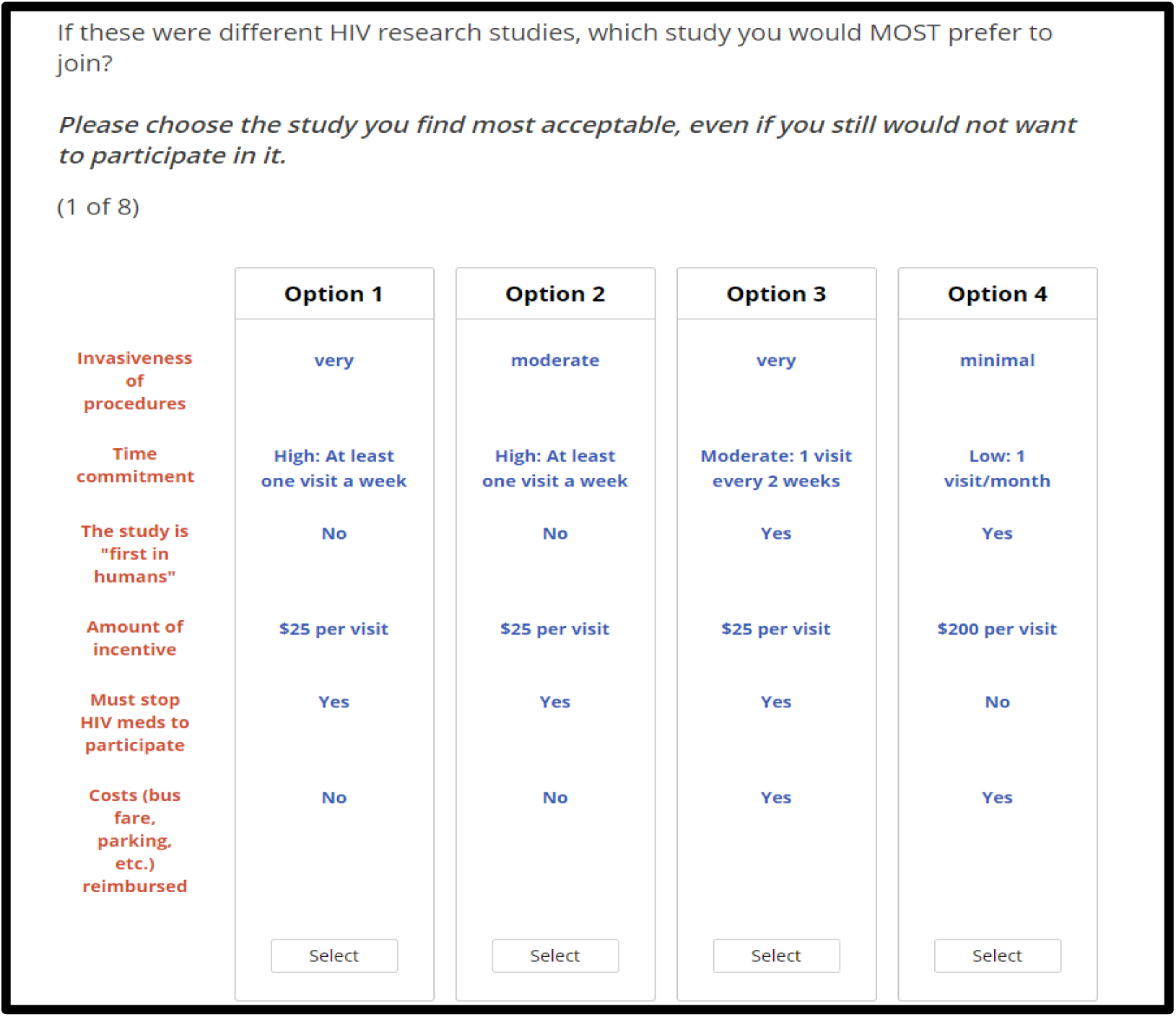
Conjoint analysis task on preferred HIV –cure-related research study scenario completed by participants (one of eight tasks)

Finally, the final attributes were each assigned 2-3 levels. Level values were determined by the study team, drawing on collective experience from reviewing or participating in HIV cure research studies, and were corroborated by the CAB for authenticity. Table 2 presents the final attribute list and its corresponding levels.

**Table 2:** Final attributes (N=6) and corresponding levels used in the CBC experiments.

| Attribute | Level 1 | Level 2 | Level 3 |
| --- | --- | --- | --- |
| **Invasiveness of procedures** | Minimal<br>*Example: only a blood draw* | Moderate<br>*Example: blood draw + a needle biopsy* | Very<br>*Example: blood draw, needle biopsies, lumbar puncture* |
| **Time commitment** | Low<br>*1 visit per month* | Moderate<br>*1 visit every 2 weeks* | High<br>*At least 1 visit per week* |
| **The study is "first in humans"** | Yes | No | (not used) |
| **Amount of incentive** | \$25 per study visit | \$100 per study visit | \$200 per study visit |
| **Must stop HIV meds to participate** | Yes | No | (not used) |
| **Costs (bus fare, parking, etc.) reimbursed** | Yes | No | (not used) |

### Data collection and analysis

The six attributes and their levels were entered into Lighthouse Studio, which applied a fractional factorial design to identify the final eight tasks for the CBC experiment. Participants were provided with a link to the CBC survey, which included a brief demographic survey. For the CBC experiment, Lighthouse Studio displayed a series of eight “tasks,” in each of which they chose their preferred HIV cure research study scenario from the four studies displayed, and, in doing so, provided information on the study attributes and levels they preferred most. The order in which the scenarios were displayed was randomized across participants. **Figure 1** displays an example of one of the conjoint tasks.

We summarized the demographics to describe the study population’s characteristics. We analyzed CBC data within Lighthouse Studio to compute the attribute utilities (the relative desirability of within-attribute levels), and computed the attribute importance of each attribute (i.e., the weight of each attribute on participants’ choices for a given hypothetical study scenario). Chi-squared tests were used to determine whether the differences in attribute-level utilities were significant at p ≤ 0.01.

## RESULTS

A total of N=88 people participated across all three groups (n=65 PWLH; n=11 HIV researchers; n=12 IRB members/bioethicists); **Table 3**. Across all participants, most were male (61%), white (51%), and educated (all had a high school education or higher).

**Table 3:**
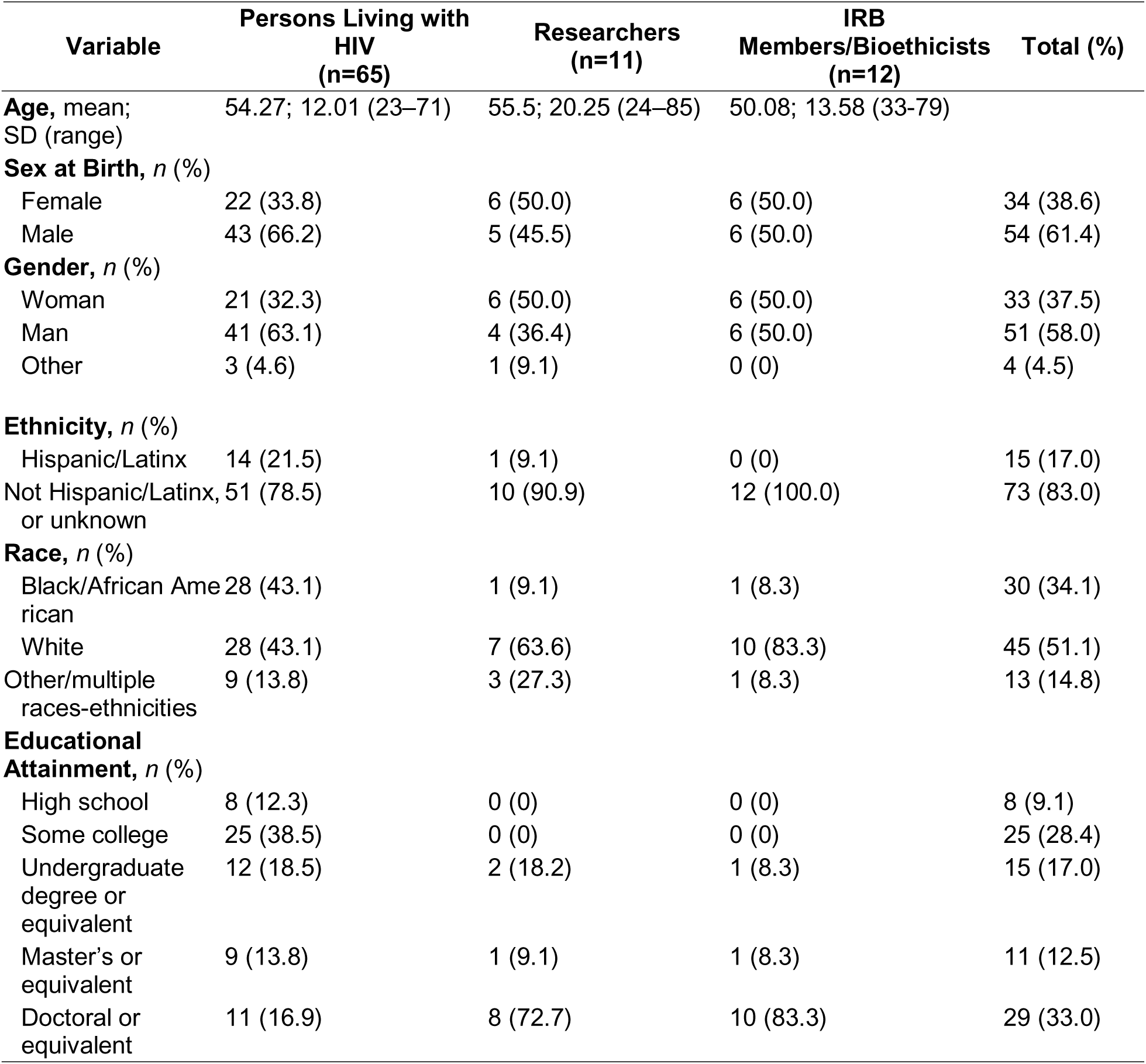
Participant characteristics (N=88)

### Attribute-level utilities

Attribute utilities, that is, the relative preference of levels *within* (but not between) each attribute, revealed a preferred level for all attributes (p<0.01) except for the “time commitment” and “cost reimbursed” attributes (**Table 4**).

**Table 4:** HIV research attribute utility preferences among N=88 respondents.

| Attribute (level options) | Level options | Most preferred level | Within-attribute Chi-Square (* = $p < 0.01$) |
| --- | --- | --- | --- |
| Invasiveness of procedures | minimal, moderate, very | minimal | **50.917\*** |
| Time Commitment | low, moderate, high | moderate | 3.345 |
| The study is “first in humans” | yes, no | no | **13.110\*** |
| Amount of incentive | \$25, \$100, \$200 (per study visit) | \$200 | **91.417\*** |
| Must stop HIV meds to participate | yes, no | no | **55.520\*** |
| Costs (bus fare, parking, etc.) reimbursed | yes, no | yes | 3.773 |

### Overall relative attribute importance – all participant types combined

The overall relative attribute importances (that is, the *between*-attribute preferences) were amount paid (29.3%); invasiveness (22.3%); must stop HIV meds to participate (17.6%); time commitment (12.5%); the study was first in humans (10.6%); cost reimbursement (7.6%).

### Attribute importance by participant type

When the attribute importance was segmented by participant type, differences were observed in the importance weight of the attributes as follows (attributes listed in order of most to least important; all attribute weights sum to 100% for each participant type): PLWH—amount paid (30.8%), invasiveness (20.2%), must stop HIV meds to participate (18.0%), time commitment (13.1%), whether first in humans (9.8%), and cost reimbursement (8.0%); Researchers—invasiveness (27.1%), amount paid (26.1%), time commitment (14.6%), must stop HIV meds to participate (13.4%), whether first in humans (10.7%), and cost reimbursement (8.2%); and, IRB members/bioethicists—invasiveness (28.2%), amount paid (25.1%), must stop HIV meds to participate (19.4%), whether first in humans (14.4%), time commitment (7.6), cost reimbursement (5.3%); **Figure 2**.

**Figure 2:**
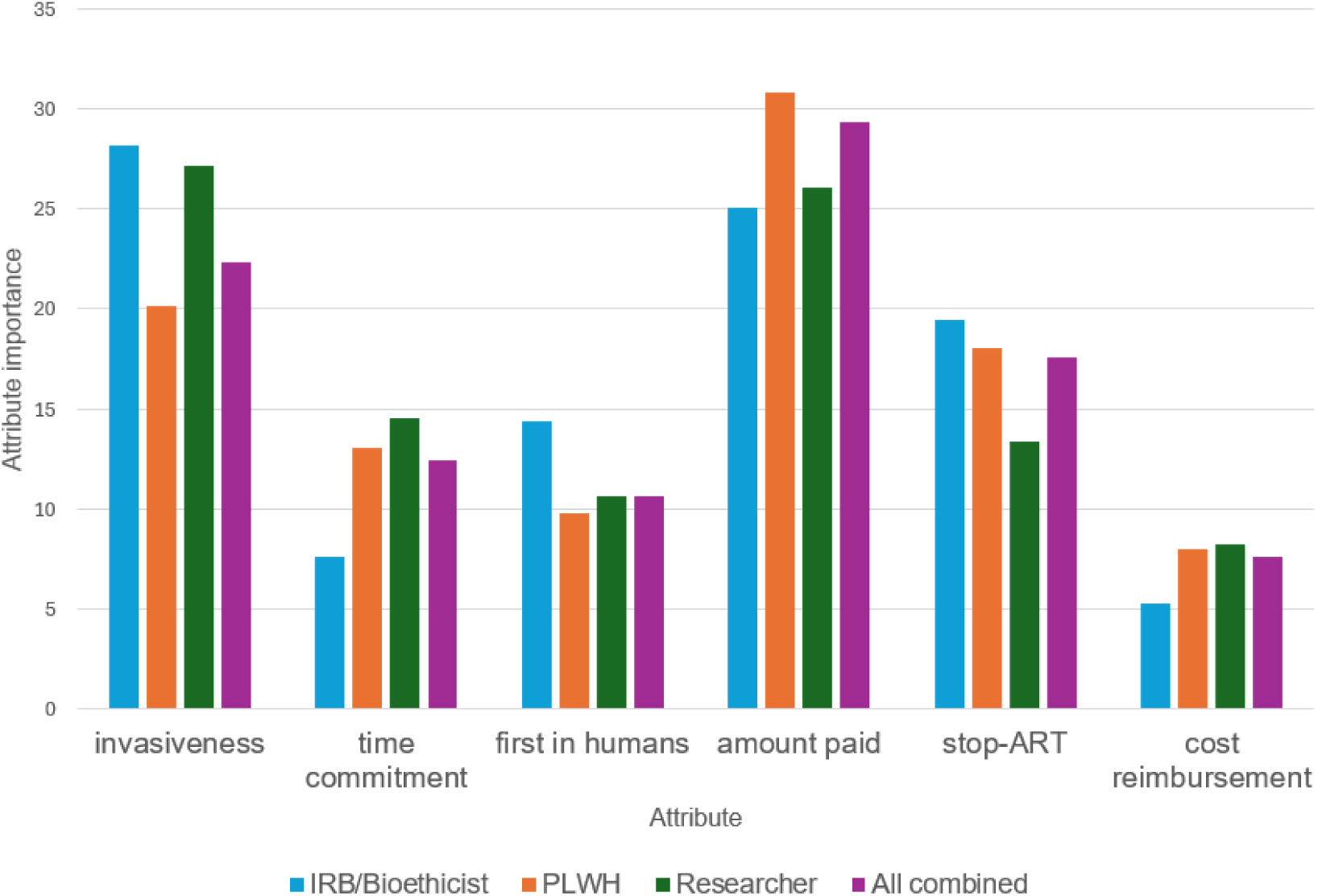
Attribute importance in hypothetical HIV study, overall and segmented by participant type (N=88)

## DISCUSSION

Overall, across all participant types (PLWH, researchers, and IRB members/bioethicists), the cash incentive amount was the primary factor driving the selection of hypothetical research scenarios. The next two most important attributes were the invasiveness of the research procedures and the need to stop HIV medication to participate in the study. However, when the CBC data were analyzed by participant type, differences in preferred attributes emerged. PLWH prioritized the participation incentive amount paid over the second most important attribute (invasiveness of the study procedures), whereas HIV researchers and IRB members/bioethicists prioritized study invasiveness over the amount paid. These findings suggest that while all stakeholder groups most prioritized similar attributes (incentive amount paid and procedure invasiveness), PLWH, who would be participating in HIV cure research trials, chose the amount paid as the factor that most influenced their choice of hypothetical HIV research trials. This finding is particularly significant because of the lack of emphasis on payment practices, including the participant payment amount, in research design.

### Incentive Payment Amount

Participant incentive payment amount was ranked as the most important attribute in a hypothetical HIV cure-related research study, compared to invasiveness, stopping HIV medication, time commitment, first-in-human studies, and ancillary cost reimbursement. Moreover, the highest incentive payment amount ($200 per study visit) was significantly preferred over lower amounts ($25 or $100 per study visit). While it is not surprising that more money is preferred over less money, it is striking that during the attribute ranking and selection phase conducted by the CAB, the amount of incentive payment provided was ranked in 9^th^ place, which may belie its true importance to prospective study participants in HIV cure research based on the CBC results. Indeed, despite its importance, there has been little research on the appropriate amount of incentive payment for research participants. In a Singapore-based study with 612 respondents from the Health Opinions Panel Singapore and the Singapore National Center for Infectious Diseases, participants ranked the importance of various factors influencing payment amounts as part of a broader study on public attitudes towards controlled human infection studies. A total of 454 (74%) respondents in that study indicated that factors such as the risk of side effects, pain, time required, the number of invasive and non-invasive investigations, and inconvenience due to the trial should all be considered when determining appropriate compensation [39]). The authors concluded that payment may not be the only reason people participate in biomedical research, but it is a major motivator for enrolling in a study. Our study corroborates the importance of incentive payments, consistent with the Singaporean study, but more data are needed to provide clearer guidance on appropriate payment amounts for research participation, especially in HIV cure-related research. Wolf and colleagues conducted a content analysis of consent forms and protocols available on clinicaltrials.gov to determine what information researchers provide regarding payment and found that, while studies often provided payment for participation, the purpose of the payment was rarely included in consent forms (i.e., compensation, reimbursement, incentive, or appreciation) [40].

### Invasiveness

In our hypothetical study scenarios, IRB members/bioethicists and researchers ranked invasiveness as the most important factor for participation in potential HIV research, in clear contrast with PLWH’s ranking of incentive payment amounts as the most important factor. A 2024 study that focused on social marketing and recruitment of PLWH reported that participants were concerned about study procedures, including investigational medications, invasive procedures, and genetic testing. Some of these concerns were rooted in participants’ demographic backgrounds. Despite hesitations, researchers in that study found that compensation influenced whether participants enrolled in clinical trials that conducted more invasive procedures [41]. Although our data showed that participants preferred minimal invasiveness, this study suggests that payment may be more important, raising concerns that riskier studies may be deemed acceptable because of the payment amount.

### Stopping HIV Medication

A requirement to temporarily stop (i.e., pause for the duration of the study) HIV medication was the third most important factor in selecting hypothetical study scenarios among all participants. When stratified by participant group, however, for IRB members/bioethicists, pausing HIV medication had a greater influence on the selection of the hypothetical scenarios compared to PLWH (the potential research participants who would be asked to discontinue HIV treatment and therefore also deal with any related side effects). These discordant findings may indicate factors beyond medical consequences, such as legal and liability issues that researchers may face if study participants experience harm from study procedures, even when participants understand the risks of participation. Research on analytical treatment interruptions among participants in HIV cure-related studies is mixed and nuanced, and dependent on the study’s specific characteristics. Studies of HIV cure research involving analytical treatment interruptions consistently show that participants exhibit conditional willingness to discontinue ART. While participants in HIV cure research are motivated by altruism and a desire to contribute to scientific advancement, including a curiosity about their own immunologic response [42], prior research reports substantial concerns regarding viral rebound, potential health harms, and risk of HIV transmission to partners, alongside psychological stress and social implications such as stigma during analytical treatment interruptions [43,44]). Preferences regarding ART discontinuation are therefore highly contingent on trial design, with participants favoring stringent safety monitoring, clear ART restart criteria, and robust support for partner protection and psychosocial well-being [42,43, 45].

### Time Commitment and Ancillary Costs

Time commitment and whether ancillary costs (bus fare, parking) were reimbursed were not statistically different in our stakeholder analysis. However, reimbursement can help reduce the burden of research participation, especially if participants must travel or miss work to participate [41]), and previous research reports that offers of payment could convince people to participate if they believe that their time (professional or private) is valuable [46]). Our study demonstrates that, while time commitment and ancillary costs play a role in study participation, these factors are secondary to payment amount, invasiveness of study procedures, and whether ART discontinuation is required as part of the design.

### Limitations

Our study had several limitations. Participants were recruited by convenience sampling and were not representative of all stakeholders in HIV cure research. Similarly, the attributes and corresponding levels could differ for certain types of HIV research, and the restriction to six attributes, necessary to prevent cognitive overload, may have inadvertently obscured the impact of other important attributes that were not included. Of course, research studies are complicated and may include more attributes than our research highlighted. Nonetheless, we followed a stepwise process to arrive at the most important attributes, which were validated by the CAB. Since this study was conducted in the United States, the findings may not be directly transferable to low- or middle-income countries or settings with different socioeconomic conditions. The ethical considerations surrounding incentives, including the potential for undue influence, may differ substantially in contexts where poverty and financial vulnerability are more prevalent.

## Conclusion

In conclusion, payment was the most important attribute across hypothetical HIV cure research studies among all participant types, and the highest payment amount was most preferred. Future research should develop realistic, yet hypothetical HIV cure-related studies based on existing research and payment practices of studies registered on ClinicalTrials.gov, and ask stakeholders for the appropriate payment amounts for those studies. We also argue that transparency of research incentives should be a common practice when disseminating research protocols and findings [47]. These data would help facilitate participant payment decision-making, including previous practices and guidance for future payment amounts.

### Public Health Implications

These findings highlight several important implications for policy and ethical oversight in HIV cure–related research. First, the strong influence of financial incentives on participation decisions underscores the need for clearer, standardized guidance on appropriate payment levels. Current practices are inconsistent and often lack transparency, suggesting that regulators and IRBs should establish more explicit frameworks for determining and reporting participant compensation. Differences across stakeholder groups, especially where people living with HIV prioritize payment more strongly than researchers and IRB members, suggest the need to better integrate participant perspectives into research design and ethical review. Greater use of community engagement mechanisms, such as community advisory boards, may help align and truthfully reflect study design with participant values while maintaining ethical standards.

The importance of study characteristics such as procedure invasiveness and antiretroviral therapy interruption indicates that policies should require enhanced protections and clearer consent processes for high-risk trial features, including robust monitoring and risk mitigation strategies. The limited reporting of payment practices highlights a broader need for greater transparency in research incentives, including disclosure of payment amounts and their intended purpose in protocols and publications. Such transparency would support ethical accountability and enable the development of evidence-based guidance for participant compensation in future research.

## Data Availability

All data produced in the present study are available upon reasonable request to the authors.

## REFERENCES

1. Arnold, M. P., Evans, D., & Vergel, N. (2015). Recruitment and ethical considerations in HIV cure trials requiring treatment interruption. Journal of Virus Eradication, 1(1), 43–48. 10.1016/s2055-6640(20)31148-1.

2. Dubé, K., Evans, D., Sylla, L., Taylor, J., Weiner, B. J., Skinner, A., Thirumurthy, H., Tucker, J. D., Rennie, S., & Greene, S. B. (2017). Willingness to participate and take risks in HIV cure research: survey results from 400 people living with HIV in the US. Journal of virus eradication, 3(1), 40–50.e21. 10.1016/S2055-6640(20)30295-8.

3. Largent E. (2017). For love and money: the need to rethink benefits in HIV cure studies. Journal of medical ethics, 43(2), 96–99. 10.1136/medethics-2015-103119.

4. Lee, R., Cui, R. R., Muessig, K. E., Thirumurthy, H., & Tucker, J. D. (2014). Incentivizing HIV/STI testing: a systematic review of the literature. AIDS and behavior, 18(5), 905–912. 10.1007/s10461-013-0588-8.

5. London AJ, Borasky DA Jr, Bhan A, for the Ethics Working Group of the HIV Prevention Trials Network (2012) Improving Ethical Review of Research Involving Incentives for Health Promotion. PLOS Medicine 9(3): e1001193. 10.1371/journal.pmed.1001193.

6. Al, P., Brehaut, J., Gillies, K., Presseau, J., Yee, M.-L., & Weijer, C. (2026). The ethical permissibility of financial incentives. *Medicine*, Health Care and Philosophy, 29(2), 535–547. 10.1007/s11019-025-10315-1.

7. Brown, J. A., Serrato, C. A., Hugh, M., Kanter, M. H., Spritzer, K. L., & Hays, R. D. (2016). Effect of a post-paid incentive on response rates to a web-based survey. Survey Practice, 9(1), 1–7. 10.29115/sp-2016-0001.

8. Grady C. (2005). Payment of clinical research subjects. The Journal of clinical investigation, 115(7), 1681–1687. 10.1172/JCI25694.

9. Webinar follow-up: compensation or inducement? What IRBs need to know about paying subjects for participation. Amp&rsand. 2017 May 8. URL: https://blog.primr.org/webinar-follow-compensation-inducement/.

10. Bentley, J. P., & Thacker, P. G. (2004). The influence of risk and monetary payment on the research participation decision making process. Journal of medical ethics, 30(3), 293–298. 10.1136/jme.2002.001594.

11. Lo, B., Grady, C., & Working Group on Ethics of the International AIDS Society (2013). Ethical considerations in HIV cure research: points to consider. Current opinion in HIV and AIDS, 8(3), 243–249. 10.1097/COH.0b013e32835ea1c5.

12. Kamuya, D. M., Marsh, V., Njuguna, P., Munywoki, P., Parker, M., & Molyneux, S. (2014). “When they see us, it’s like they have seen the benefits!”: experiences of study benefits negotiations in community-based studies on the Kenyan Coast. BMC medical ethics, 15, 90. 10.1186/1472-6939-15-90.

13. Dubé, K., Henderson, G. E., & Margolis, D. M. (2014). Framing expectations in early HIV cure research. Trends in microbiology, 22(10), 547–549. 10.1016/j.tim.2014.08.003.

14. Resnik D. B. (2015). Bioethical Issues in Providing Financial Incentives to Research Participants. Medicolegal and bioethics, 5, 35–41. 10.2147/MB.S70416.

15. Payment and reimbursement to research subjects. U.S. Food & Drug Administration. 2018 Jan 25. https://www.fda.gov/regulatory-information/search-fda-guidance-documents/payment-and-reimbursement-research-subjects.

16. Department of Health, Education, and Welfare, & National Commission for the Protection of Human Subjects of Biomedical and Behavioral Research (2014). The Belmont Report. Ethical principles and guidelines for the protection of human subjects of research. The Journal of the American College of Dentists, 81(3), 4–13.

17. Greene, K. Y., Brown, B., Galea, J. T., Lopez, J. L., Dubé, K., Taylor, J., Christensen, C., & Polonijo, A. N. (2026). Determining Ethical Incentives in Biomedical HIV Research: Insights from Researchers, Regulators, and People Living with HIV. Journal of empirical research on human research ethics: JERHRE, 15562646261441345. Advance online publication. 10.1177/15562646261441345.

18. Polonijo, A. N., Dubé, K., Galea, J. T., Greene, K. Y., Taylor, J., Christensen, C., & Brown, B. (2022). Attitudes Toward Payment for Research Participation: Results from a U.S. Survey of People Living with HIV. AIDS and behavior, 26(10), 3267–3278. 10.1007/s10461-022-03660-2.

19. Groth S. W. (2010). Honorarium or coercion: use of incentives for participants in clinical research. *The Journal of the New York State Nurses’* Association, 41(1), 11–22.

20. Cryder, C. E., John London, A., Volpp, K. G., & Loewenstein, G. (2010). Informative inducement: study payment as a signal of risk. Social science & medicine (1982), 70(3), 455–464. 10.1016/j.socscimed.2009.10.047.

21. Walker RL, MacKay D, Waltz M, Lyerly AD, Fisher JA. Ethical Criteria for Improved Human Subject Protections in Phase I Healthy Volunteer Trials. Ethics Hum Res. 2022;44(5):2–21. doi:10.1002/eahr.500139

22. Ripley E. B. (2006). A Review of Paying Research Participants: It’s Time to Move Beyond the ethical Debate. Journal of empirical research on human research ethics: JERHRE, 1(4), 9–20. 10.1525/jer.2006.1.4.9

23. Brown, B., Marg, L., Michels, E., Zhang, Z., Kuzmanović, D., Dubé, K., & Galea, J. T. (2021). Comparing Payments Between Sociobehavioral and Biomedical Studies in a Large Research University in Southern California. Journal of empirical research on human research ethics: JERHRE, 16(1-2), 117–124. 10.1177/1556264620987773.

24. Gelinas, L., Largent, E. A., Cohen, I. G., Kornetsky, S., Bierer, B. E., & Fernandez Lynch, H. (2018). A Framework for Ethical Payment to Research Participants. The New England journal of medicine, 378(8), 766–771. 10.1056/NEJMsb1710591.

25. Largent, E. A., Walter, S., & Edelmayer, R. M. (2022). Pgarticipant first: Improving communication with and support for participants in trials that end early. Alzheimer’s & Dementia, 18(S10). 10.1002/alz.061292.

26. Evans D. (2017). An activist’s argument that participant values should guide risk-benefit ratio calculations in HIV cure research. Journal of medical ethics, 43(2), 100–103. 10.1136/medethics-2015-103120.

27. Saberi, P., Campbell, C. K., Sauceda, J. A., Ndukwe, S., & Dubé, K. (2022). Perceptions of Risks and Benefits of Participating in HIV Cure-Related Research Among Diverse Young Adults Living with HIV in the United States: Qualitative Research Findings. AIDS research and human retroviruses, 38(8), 649–659. 10.1089/AID.2021.0192.

28. Brown, B., & Galea, J. T. (2019). The effect of incentives on HIV research must be studied. The lancet. HIV, 6(3), e142–e143. 10.1016/S2352-3018(18)30357-6.

29. Trung LQ, Morra ME, Truong ND, et al. A systematic review finds underreporting of ethics approval, informed consent, and incentives in clinical trials. J Clin Epidemiol. 2017;91:80–86. doi:10.1016/j.jclinepi.2017.08.007

30. Galea JT, Greene KY, Nguyen B, et al. Evaluating the Impact of Incentives on Clinical Trial Participation: Protocol for a Mixed Methods, Community-Engaged Study. JMIR Res Protoc. 2021;10(11):e33608. Published 2021 Nov 23. doi:10.2196/33608

31. Green, P. E., & Srinivasan, V. (1990). Conjoint Analysis in Marketing: New Developments with Implications for Research and Practice. Journal of Marketing, 54(4), 3–19. 10.2307/1251756

32. de Bekker-Grob, E. W., Donkers, B., Bliemer, M. C. J., Veldwijk, J., & Swait, J. D. (2020). Can healthcare choice be predicted using stated preference data?. Social science & medicine (1982), 246, 112736. 10.1016/j.socscimed.2019.112736.

33. de Bekker-Grob, E. W., Swait, J. D., Kassahun, H. T., Bliemer, M. C. J., Jonker, M. F., Veldwijk, J., Cong, K., Rose, J. M., & Donkers, B. (2019). Are Healthcare Choices Predictable? The Impact of Discrete Choice Experiment Designs and Models. Value in health: the journal of the International Society for Pharmacoeconomics and Outcomes Research, 22(9), 1050–1062. 10.1016/j.jval.2019.04.1924.

34. Quaife, M., Terris-Prestholt, F., Di Tanna, G. L., & Vickerman, P. (2018). How well do discrete choice experiments predict health choices? A systematic review and meta-analysis of external validity. The European journal of health economics: HEPAC: health economics in prevention and care, 19(8), 1053–1066. 10.1007/s10198-018-0954-6.

35. Beusterien, K. M., Dziekan, K., Flood, E., Harding, G., & Jordan, J. C. (2005). Understanding patient preferences for HIV medications using adaptive conjoint analysis: Feasibility Assessment. Value in Health, 8(4), 453–461. 10.1111/j.1524-4733.2005.00036.x.

36. Tang, E. C., Galea, J. T., Kinsler, J. J., Gonzales, P., Sobieszczyk, M. E., Sanchez, J., & Lama, J. R. (2016). Using conjoint analysis to determine the impact of product and user characteristics on acceptability of rectal microbicides for HIV prevention among Peruvian men who have sex with men. Sexually transmitted infections, 92(3), 200–205. 10.1136/sextrans-2015-052028.

37. Kinsler, J. J., Cunningham, W. E., Nureña, C. R., Nadjat-Haiem, C., Grinsztejn, B., Casapia, M., Montoya-Herrera, O., Sánchez, J., & Galea, J. T. (2012). Using conjoint analysis to measure the acceptability of rectal microbicides among men who have sex with men in four South American cities. AIDS and behavior, 16(6), 1436–1447. 10.1007/s10461-011-0045-5.

38. Orme B. Getting started with conjoint analysis: strategies for product design and pricing research. 2nd ed. Madison: Research Publishers LLC; 2010.

39. Young, B., Giubilini, A., Sam, X. H., Lysaght, T., Tambyah, P. A., Schaefer, G. O., & Savulescu, J. (2025). An Ethical Analysis of Public Attitudes towards Controlled Human Infection Studies in Singapore: Acceptability and Payment. Asian bioethics review, 18(1), 117–134. 10.1007/s41649-024-00335-z

40. Wolf LE, Kench S, Ledford CJW. A taxing problem: The impacts of research payment practices on participants and inclusive research. PLoS One. 2024;19(6):e0303112. Published 2024 Jun 6. doi:10.1371/journal.pone.0303112

41. Idnay, B., Cordoba, E., Ramirez, S. O., Xiao, E., Wood, O. R., Batey, D. S., Garofalo, R., & Schnall, R. (2024). Social Marketing Perspective on Participant Recruitment in Informatics-Based Intervention Studies. AIDS and behavior, 28(9), 2836–2849. 10.1007/s10461-024-04355-6.

42. Nguyen, E., Korolkova, A., Ahmed, A., Meanley, S., Dee, L., Hussain, M., Wan, F., Hoh, R., Rodriguez, A., Figueroa, T., Cohn, L. B., Deeks, S. G., Peluso, M. J., Eskaf, S., Sugarman, J., Sauceda, J. A., & Dubé, K. (2025). Participant Perspectives in an HIV Treatment Interruption Study in San Francisco, United States. AIDS research and human retroviruses, 41(11), 505–516. 10.1177/08892229251375470.

43. Lau, J. S. Y., Smith, M. Z., Allan, B., Martinez, C., Power, J., Lewin, S. R., & McMahon, J. H. (2020). Perspectives on Analytical Treatment Interruptions in People Living with HIV and Their Health Care Providers in the Landscape of HIV Cure-Focused Studies. AIDS research and human retroviruses, 36(4), 260–267. 10.1089/AID.2019.0118.

44. Ndung’u, T., Dong, K. L., Colby, D. J., Deeks, S. G., Cotton, M. F., Dubé, K., Goulder, P. J., Julg, B., Nsubuga, M., Ajibola, G., Bar, K. J., Bengu, N., Bwakura-Dangarembizi, M., Caskey, M., Cromhout, G., Kekitiinwa, A. R., Kilembe, W., Kuhn, L., Likhitwonnawut, U., … Trautmann, L. (2026). Recommendations from the 2nd Consensus Workshop on Analytical Treatment Interruption in HIV research trials. The Lancet HIV, 13(4). 10.1016/s2352-3018(25)00373-x.

45. Sauceda JA, Lisha N, Ahmed A, et al. Anxiety Symptom Ratings Before and During the Interruption of Antiretroviral Therapy as Part of HIV Cure-related Studies. J Acquir Immune Defic Syndr. 2026;101(5):496–502. doi:10.1097/QAI.0000000000003834

46. Różyńska J. (2022). The ethical anatomy of payment for research participants. Medicine, health care, and philosophy, 25(3), 449–464. 10.1007/s11019-022-10092-1.

47. Brown, B., Galea, J. T., Davidson, P., & Khoshnood, K. (2016). Transparency of participant incentives in HIV research. The Lancet HIV, 3(10). 10.1016/s2352-3018(16)30150-3.

